# Improving Power and Accuracy in Randomized Controlled Trials of Pain Treatments by Accounting for Concurrent Analgesic Use

**DOI:** 10.1101/2021.10.30.21265709

**Authors:** Pradeep Suri, Patrick J. Heagerty, Anna Korpak, Mark P. Jensen, Laura S. Gold, Kwun C. G. Chan, Andrew Timmons, Janna Friedly, Jeffrey G. Jarvik, Aaron Baraff

## Abstract

The 0 to 10 numeric rating scale (NRS) of pain intensity is a standard outcome in randomized controlled trials (RCTs) of pain treatments. For individuals taking analgesics, there may be a disparity between “observed” pain intensity (the NRS, irrespective of concurrent analgesic use), and “underlying” pain intensity (what the NRS would be had concurrent analgesics not been taken). Using a contemporary causal inference framework, we compare analytic methods that can potentially account for concurrent analgesic use, first in statistical simulations, and second in analyses of real (non-simulated) data from an RCT of lumbar epidural steroid injections (LESI). The default analytic method was ignoring analgesic use, which is the most common approach in pain RCTs. Compared to ignoring analgesic use and other analytic methods, simulations showed that a quantitative pain and analgesia composite outcome based on adding 1.5 points to observed pain intensity for those who were taking an analgesic (the QPAC_1.5_) optimized power and minimized bias. Analyses of real RCT data supported the results of the simulations, showing greater power with analysis of the QPAC_1.5_ as compared to ignoring analgesic use and most other methods examined. We propose alternative methods that should be considered in the analysis of pain RCTs.

## INTRODUCTION

Pain is a major public health problem that affects 100 million adults in the United States (US) and costs up to $635 billion annually.^1,2^ More effective treatments for pain are needed.^3^ A potential obstacle to the development of new pain treatments is that conventional methods for analyzing pain intensity outcomes in randomized controlled trials (RCTs) of pain treatments for acute and chronic pain do not account for concurrent analgesic use.

One of the most common measures of pain intensity in pain RCTs and clinical care is the 0-10 Numeric Rating Scale (NRS).^4^ Most non-surgical pain treatments used in outpatient settings have small treatment effects on pain, corresponding to 0.5 to 1.0-point improvements in the NRS without correcting for analgesic use.^5-7^ However, as many as 72-80% of patients with pain use “over-the-counter” or prescribed analgesics, self-regulating analgesic use to achieve tolerable pain levels.^8,9^ Given that the average treatment effects of analgesics in RCTs are also of 0.5 to 1.0-NRS-point improvements,^5,10,11^ the effect size of concurrent analgesic use may equal or exceed the effect size of the pain treatment under study in an RCT. While randomization in an RCT can balance treatment and control groups with respect to many characteristics, including concurrent analgesic use at baseline, differential post-randomization analgesic use between treatment and control groups may occur when a treatment is effective and can mask important differences in underlying pain. As a result, clinically relevant effects of a treatment in a pain RCT may be obscured, attenuating estimates of treatment effects and decreasing statistical power.

A number of analytic methods have been proposed to minimize imprecision or bias due to concurrent medication use in the cardiovascular literature.^12-15^ However, to date, these methods have largely not been used to account for analgesic use in studies of pain, and it is unknown whether they are applicable to pain treatment RCTs. The overarching aim of the current study was to begin to address this critical knowledge gap. First, we formalized the estimation goals (estimands) of pain RCTs through specification of a causal framework that considers analgesic use. Next, we conducted a simulation study to compare different methods of accounting for concurrent analgesic use in the context of a hypothetical RCT designed to examine the effect of a pain treatment on pain intensity. The goals of the simulations were to compare these analytic methods with regards to (1) statistical power to detect a treatment effect, and (2) bias of estimated treatment effects. Last, we analyzed “real” (non-simulated) patient data from a pain treatment RCT to compare these analytic methods with regards to statistical power.

## METHODS

### Overview of concepts and terminology

A conceptual model of how pain treatments may affect a patient’s pain intensity and analgesic use is presented in **Figure 1**. Conventional intent-to-treat (ITT) analyses as typically conducted in RCTs of pain treatments do not account for analgesic use, and therefore estimate the overall effect (total effect) of a treatment on “*observed”* pain intensity: current pain as reported by the patient, irrespective of whether patients are using analgesics concurrently. Such “total effects” do not account for post-randomization between-group differences in analgesic use. Specifically, analyses of total effects in pain RCTs treat as equivalent an individual who achieves a tolerable pain intensity (e.g., NRS=4) at follow-up without concurrent analgesic use and another individual who achieves the same pain intensity at follow-up (NRS=4) but uses analgesics. However, researchers and clinicians likely perceive these two individuals as being different, with the second having achieved a less optimal pain state. Consistent with this, the conceptual model presented in **Figure 1** assumes that the effect of greatest interest-what patients and clinicians really care about-is that of a treatment on “*underlying*” pain intensity, reflecting the hypothetical situation of what a patient’s pain intensity would be were they not using analgesics concurrently.

**Figure 1.**
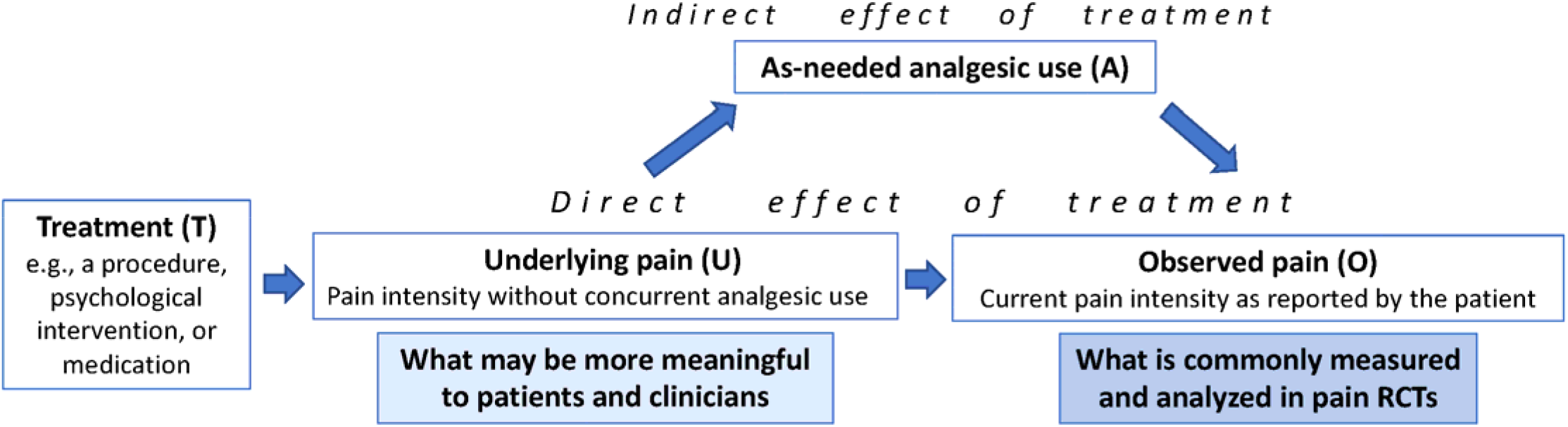
Conceptual model of the effects of a pain treatment on underlying pain and observed pain.

In common epidemiologic terminology, the “total effect” of a treatment on observed pain intensity as typically estimated in ITT analyses that do not account for concurrent analgesic use is a combination of the “direct effect” of the pain treatment on underlying pain (U) and observed pain intensity (O), and an “indirect effect” on observed pain intensity that is mediated through concurrent analgesic use (A), such as patients’ efforts to change their analgesic use in response to their current pain intensity (**Figure 1**).^16^. Contrary to common arguments for the use of ITT, the direct effect and the indirect effect are likely to have opposite signs or directions of effect, because individuals with higher underlying pain are more likely to use analgesics,^8^ resulting in a lower observed pain. Therefore, the ITT “total effect” when not accounting for concurrent analgesic use is likely to be attenuated towards 0. The effect of a pain treatment (T) on underlying pain intensity (U) is a “controlled direct effect,”^16^ reflecting the average difference in pain between treated and untreated patients, if all patients did not take analgesics. A narrow focus on the total effect without attempts to also characterize direct effects on pain – such as the controlled direct effect – may obscure relevant effects of the pain treatments under study.

Causal inference formalization adopts the following “potential” (counterfactual) outcomes for each patient (or research participant): *potential analgesic use* A(t), which is analgesic use should the patient be given treatment “t” (t=1 for treated and t=0 for controls); and *potential observed pain* O(t,a), should the treatment be set to “t” and analgesic use be set to “a”. For instance, O(1,0) is the value of observed pain when the patient is treated but does not use analgesics. The “controlled direct effect”- or the effect on underlying pain- can therefore be explicitly defined as the mean of O(0, 0) as compared to the mean of O(1, 0), or the difference in the mean outcomes between the control and treatment groups if you could set analgesic use (A) to 0 for all patients. We propose that the controlled direct effect of treatment- the effect of treatment on underlying pain- is the underlying construct or “estimand” of greatest relevance to patients and clinicians who are interpreting the findings of a pain RCT. We note that the term “underlying pain” used in this manuscript is <u>not</u> meant to represent an inferred latent variable such as is used in factor analysis or structural equation modeling.

### Overview of methods

As noted previously, here we conducted analyses of two types of data to address the study aims: analyses of simulated RCT data and analyses of real (non-simulated) RCT data. Statistical simulations involve generating data using pseudo-random sampling from known probability distributions, and analyzing the data generated.^17^ Simulations to examine new biostatistical methods are typically informed by summary statistics reflecting real patient data (group means, standard deviations, etc.) which allows them to closely resemble real datasets. The key strength of simulation studies over analyses of real RCT data is the ability of simulations to incorporate basic “truths” into the data to be analyzed,^17^ which are parameters of interest, such as the effect size of a pain treatment. This cannot be done with real RCT data, where the underlying truth regarding parameters of interest, such as a treatment effect, are unknown (in fact, estimating such parameters is usually the purpose of an RCT). Accordingly, simulations can provide information about potential *bias* in estimated treatment effects, which cannot be learned from analyses of real RCTs. The current study first analyzes simulated RCT data to make inferences about power and bias. We then analyze real RCT data to make inferences about power. The format of analyzing simulated data, followed by analyses of real RCT, is a standard approach for demonstrating the value of new statistical methods.^17^

In this document, to make the terminology used more easily understood by a clinical audience, we deviate from some conventions of typical causal inference notation (e.g., “Z” denoting treatment); instead, the notation used here by default takes the first letter of the construct being referred to (e.g. “T” for treatment; see **Box**).

#### Box Terminology used in models

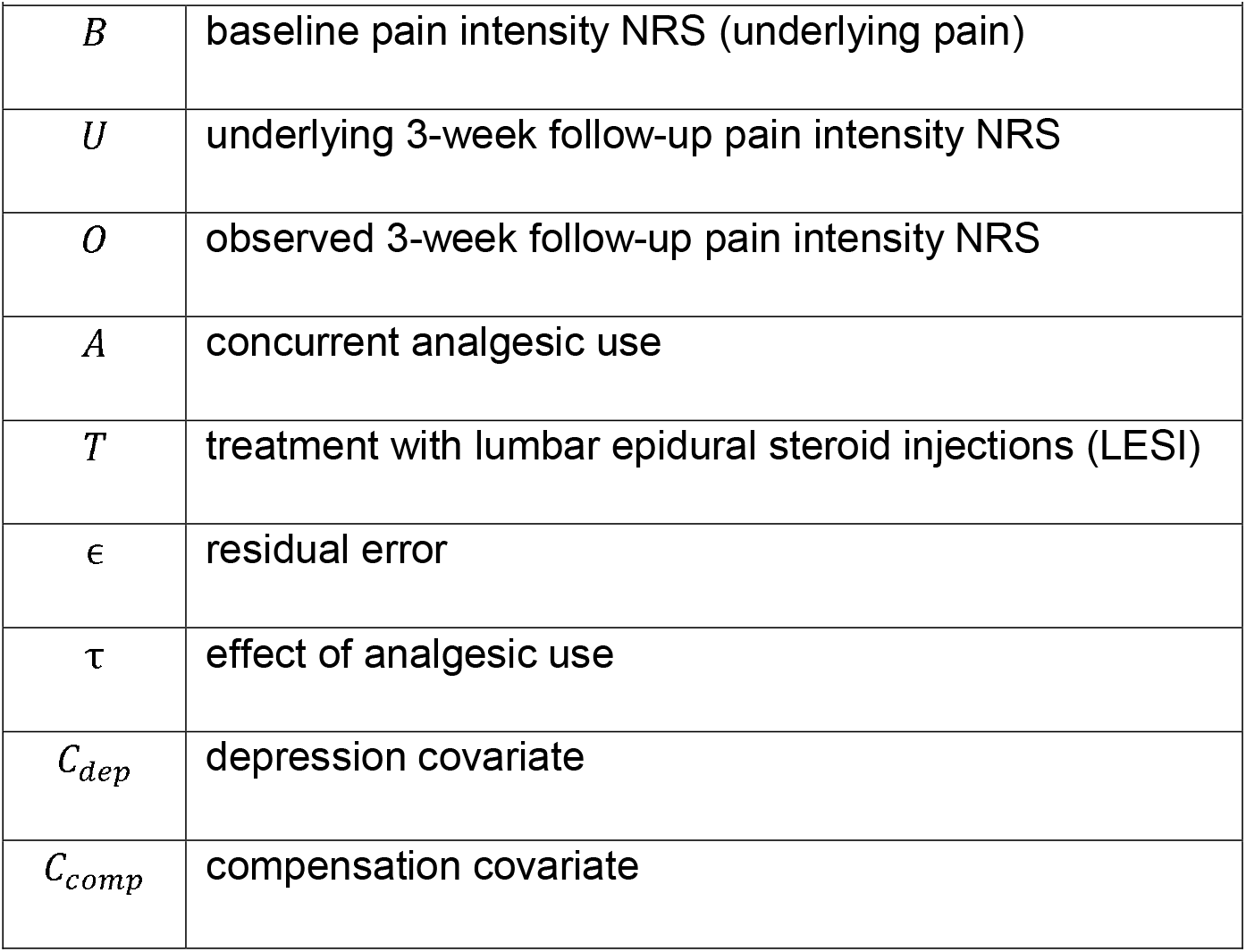

| **Box.** Terminology used in models |  |
| --- | --- |
| $B$ | baseline pain intensity NRS (underlying pain) |
| $U$ | underlying 3-week follow-up pain intensity NRS |
| $O$ | observed 3-week follow-up pain intensity NRS |
| $A$ | concurrent analgesic use |
| $T$ | treatment with lumbar epidural steroid injections (LESI) |
| $\epsilon$ | residual error |
| $\tau$ | effect of analgesic use |
| $C_{dep}$ | depression covariate |
| $C_{comp}$ | compensation covariate |

### Statistical simulations and sources of summary data

We simulated the effectiveness of lumbar epidural steroid injections (LESI), a commonly used procedure for improving pain intensity in patients with lumbar spinal stenosis (LSS), as a representative treatment for a specific pain condition.. The selection of LESI as the treatment of interest was motivated by the reasoning that the measurement of pain intensity in studies of interventional pain treatments may be particularly susceptible to influence by differential changes in analgesic use between treatment groups early in the post-randomization period, due to the early onset of action expected with LESI, and because non-steroidal anti-inflammatory drugs (NSAIDs)- a common analgesic class- are often withheld prior to LESI. Of note, between-group differences in concurrent analgesic use at baseline (*pre*-randomization) will typically be addressed by the randomization, but between-group differential concurrent analgesic *post*-randomization can still occur as a consequence of the randomized treatments; the methods studied here seek to address this problem. First, we simulated datasets (“data generation”) based on estimates of pain treatment effects from published and unpublished summary data sources, in the context of four “scenarios” reflecting different underlying assumptions, with higher scenario numbers reflecting greater complexity of the assumptions involved. Data generation was conducted using models 1-5 (described below). Second, we applied 8 analytic methods (methods A-H) to analyze the simulated datasets. For most methods, one of the linear regression “estimation models” (Models 6 and 7 below) was fit on the simulated data; these are labeled as “estimation” models to distinguish them from the data generation models (models 1-5).

First, we simulated datasets based on three sources of summary data.^18-20^ We estimated underlying pain intensity levels among those receiving LESI and the effects of LESI on pain intensity using published data from the Lumbar Epidural Steroid Injections for Spinal Stenosis (LESS) RCT.^18^ LESS was a double-blind multisite RCT which randomly assigned 400 patients with symptomatic lumbar spinal stenosis to receive LESI (n=200) or to receive control epidural injections of lidocaine alone (n=200). Baseline pain intensity in the LESS trial was 7.2 ± 1.8 NRS points, and the effect of LESI on pain intensity (vs. control) at 3-week follow-up in LESS was - 0.6 NRS points (95% confidence interval [CI] -1.2 to -0.1).^18^ We estimated the frequency of analgesic use and associations with underlying pain intensity using unpublished summary data from a recent cohort study of Veterans with low back pain, described by Kneeman et al.^19^ This cohort included 774 adults with low back pain (LBP), in which 57% of participants reported current use of analgesics. Participants were asked to report their average pain intensity over the past 2 weeks using an 11-point NRS scale; these values were considered to reflect the conventional *observed* pain intensity NRS. Separately, participants were asked about current analgesic use, and those with current analgesic use were then asked to report what their average back pain intensity in the past 2 weeks <u>would have been</u> if they were <u>not</u> taking these analgesic medications, using an 11-point NRS scale; these values were considered to reflect *underlying* pain intensity among participants with concurrent analgesic use. Mean underlying pain intensity ratings among those with concurrent analgesic use were ∼1.5 NRS points higher than observed pain intensity ratings; underlying pain intensity was assumed to be equal to observed pain intensity among those not taking analgesics. We estimated the effects of baseline covariates on pain intensity at follow-up (baseline pain intensity, receiving disability compensation, and depression) using published data from a large cohort study of 815 individuals with LBP by Pinto et al.^20^ Further descriptions of these 3 studies have been previously reported,^18,20,21^.

In order to reflect different possible relationships between the LESI treatment, underlying pain intensity, concurrent analgesic use, and covariates, we generated 4 different simulation scenarios (**Figure 2**). Scenarios 1 and 2 reflect simple relationships between treatment, underlying pain intensity, concurrent analgesic use, and covariates, with all assumptions informed by specific estimates from the summary data sources described above.^1-3^ Scenarios 3 and 4 involve more complex theoretical relationships between the LESI treatment, underlying pain intensity, concurrent analgesic use, and covariates; for Scenarios 3 and 4, where there was a lack of prior data to inform estimates for these underlying relationships, plausible small-to-moderate magnitude effect sizes were assumed. The focus in Scenarios 3 and 4 is not the specific covariates mentioned, nor their direction or magnitude. Instead, the intent is to create a more complex background of underlying relationships sufficient to challenge the analytic methods under study and illustrate their performance in different contexts. In the descriptions that follow, all effect estimates used for simulation purposes were informed by actual summary data from the 3 sources cited above,^1-3^ except for specific instances in which we state that clinically plausible (yet arbitrary) values were assumed. Broadly speaking, in Scenarios 1-3, there are two ways in which treatment may affect observed pain: (i) directly through its treatment effect on underlying pain, and (ii) indirectly by lowering underlying pain and therefore lowering the probability of analgesic use. In Scenario 4, there is an additional way in which treatment may affect observed pain: (iii) by directly lowering the probability of analgesic use in the analgesic use model, independent of (i) and (ii). Scenario 4 was motivated by multimodal or behavioral pain treatments (such as cognitive behavioral therapy) which might act in this way, with a behavioral component of treatment that aims to alter patterns of analgesic use (such as opioid use), and other aspects of treatment that aim to decrease pain.

**Figure 2.**
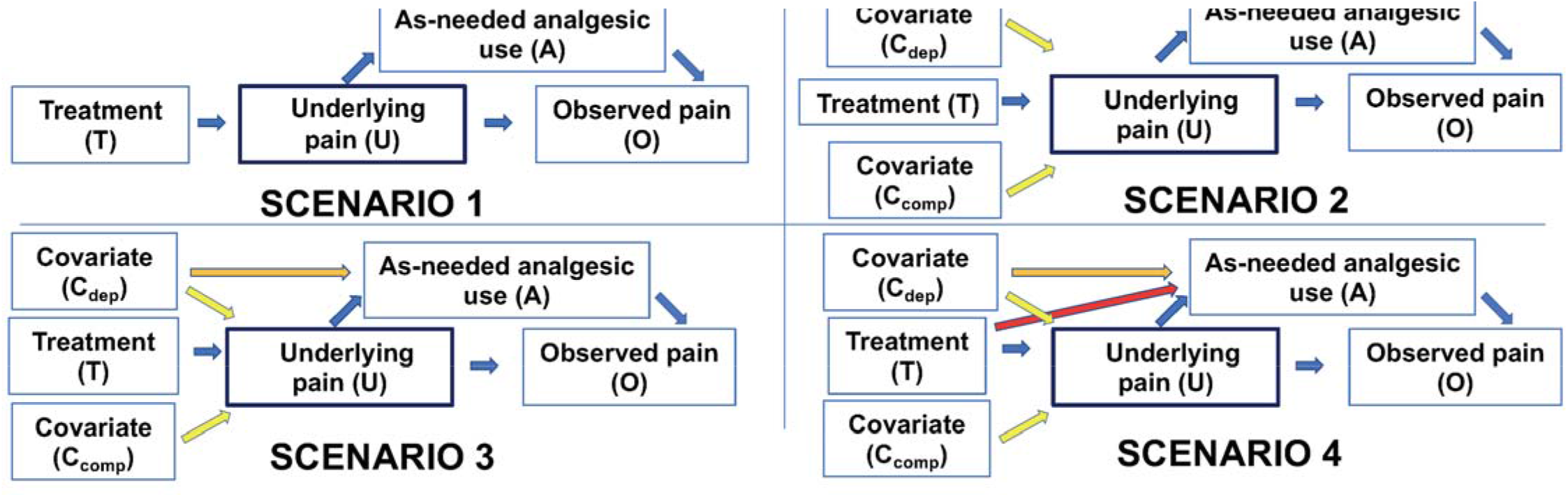
Causal diagrams representing theorized relationships between the LESI treatment, pain intensity, concurrent analgesic use, and covariates, in the simulated Scenarios. LESI=lumbar epidural steroid injection, T= treatment with lumbar epidural steroid injections (LESI), U=underlying 3-week follow-up pain intensity NRS, O=observed 3-week follow-up pain intensity NRS, A= concurrent analgesic use, C_dep_=depression covariate, C_comp_= compensation covariate

For each scenario, 10,000 simulated datasets were generated from a multipart model with underlying pain, probability of analgesic use, and observed pain after analgesic use each successively and separately simulated for 400 patients (200 per arm), emulating the sample size and design of the LESS RCT.^18^ While treatment and other covariates were assumed to have their effects on the underlying pain values, it was the observed pain values that were analyzed. For all simulation scenarios, underlying baseline pain values were assigned to each patient from a normal distribution (mean 7.2 and standard deviation 1.8), with 3-week follow-up pain NRS values derived from the LESS RCT results.^1^ All pain values were constrained to between 0 and 10, rounded to the nearest whole number. Henceforth, underlying baseline pain NRS values will be denoted by *B*, underlying pain NRS values at 3-week follow-up will be denoted by *U* and observed pain NRS values at follow-up by *O* (see Box). Note that *O*_*i*_ = *U*_*i*_ for all patients *i* = 1,…,*n* without analgesic use. Briefly, in Scenario 1 (**Figure 2**), only the treatment effect of LESI, underlying pain, observed pain, and analgesic use are modeled. Scenario 2 is similar to Scenario 1, but adds two baseline covariates commonly associated with pain-related outcomes: depression and receiving disability compensation. Scenario 3 builds upon Scenario 2 by allowing depression to affect not only underlying follow-up pain but also to affect analgesic use (i.e. depression is a confounder of the effect of analgesic on follow-up pain) (**Figure 2**). Scenario 4 further builds on Scenario 3 by also allowing the treatment variable to affect analgesic use, independent of the treatment’s effect on pain intensity.

### Scenario 1 (no covariates)

In Scenario 1, only the treatment effect for LESI was included in the model, without covariates (**Figure 2**). Values for underlying pain NRS at 3-week follow-up were generated as in Model 1,

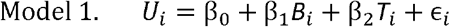

with β_0_ = 0, β_1_ = 0.5, β_2_ = − 0.6, and ∈ ∼ N(0,1.4^2^). These values were informed by the LESS results, and correspond to an approximately halving of pain intensity by 3-week follow-up, with an additional reduction of 0.6 NRS points for those receiving LESI treatment.^1^ However, a smaller value for the standard deviation of the residuals (1.4) was used in order to calibrate simulated power values to the commonly used threshold of 80%.

Analgesic use in this scenario was assumed to depend only on underlying pain values. An indicator for analgesic use (A) was assigned randomly according to a Bernoulli distribution with probability parameterized in the log-odds, or logit, using Model 2

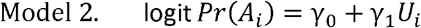

with γ_0_ = − 1.5 and γ_1_ = 0.5, informed by data from the cohort study by Kneeman et al.^4^ This corresponds to a 53.7% probability of analgesic use for the average patient, with a 1.2% increase in use for each 0.1 point increase in underlying pain.^2^

Observed pain was then derived from underlying pain by subtracting the simulated analgesic effect τ according to Model 3

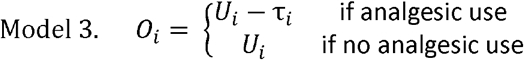

with τ ∼ N(1.5,1.4^2^), truncated at 0.^2^ Here, the mean analgesic effect was the same for all patients, not dependent on treatment, underlying pain, or any other covariates.

### Scenario 2 (also including covariates associated with underlying pain)

Scenario 2 added two baseline covariates commonly associated with pain-related outcomes: depression and receiving disability compensation (**Figure 2**). Both covariates were assigned at random from a Bernoulli distribution with probability 0.16 for depression and 0.12 for compensation, reflecting the baseline prevalence of depression and compensation from Pinto et al.^3^ Instead of Model 1, underlying follow-up pain NRS values were generated using Model 4

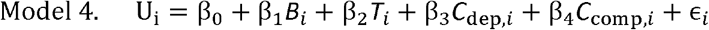

with β_0_ = 0, β_1_ = 0.5, β_2_ = − 0.6, β_3_ = 0.6, β_4_ = 0.6, and ∈ ∼ N(0,1.4^2^). These values were informed by LESS^1^ and Pinto et al. ^3^, and reflect a mean follow-up pain NRS that is 0.6 points higher for depressed patients and 0.6 NRS points higher for patients receiving disability compensation.^3^ Models for analgesic use (Model 2) and observed pain (Model 3) were the same as in Scenario 1.

### Scenario 3 (also including covariates associated with analgesic use)

Scenario 3 built upon Scenario 2 by allowing the depression covariate to affect not only underlying follow-up pain but also the probability of analgesic use and size of analgesic effect (i.e. depression as a confounder of the effect of analgesic on follow-up pain) (**Figure 2**). Due to a paucity of published data regarding predictors of actual analgesic use in LBP, a plausible and moderate magnitude effect size of depression on analgesic use (−1.0) was assumed. Instead of Model 2, the analgesic use indicator was assigned randomly according to a Bernoulli distribution with probability parameterized in the log-odds, or logit, using Model 5

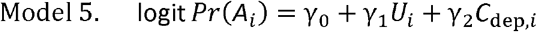

with γ_0_ = − 1.5, γ_1_ = 0.5, and γ_2_ = − 1.0. These values correspond to an additional 23.8% decrease in analgesic use for the average depressed patient. The model for underlying pain (Model 4) was the same as in Scenario 2, and observed pain was derived from underlying pain by subtracting the simulated analgesic effect τ according to Model 3, with τ_i_ ∼ N(1.5 − 0.5*C*_dep,*i*_, 1.4^2^), reflecting a mean analgesic effect that is 0.5 NRS points lower for depressed patients.

### Scenario 4 (treatment affects analgesic use)

Scenario 4 further built Scenario 3 by also allowing the treatment variable to affect the probability of analgesic use, independent of the treatment’s effect on pain intensity (**Figure 2**). This Scenario was motivated by multimodal or behavioral treatments which might act in this way, such as where a behavioral component of treatment aims to decrease analgesic use (such as opioids), and other aspects of the treatment aim to decrease pain. For this Scenario where prior data to inform treatment effects were unavailable, a plausible and small magnitude direct effect on analgesic use (−0.5) was assumed. The analgesic use indicator was assigned randomly according to a Bernoulli distribution with probability parameterized in the log-odds, or logit, using the model

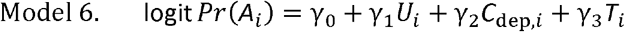

with γ_0_ = −1.5, γ_1_ = 0.5, γ_2_ = −1.0, and γ_3_ = −0.5. These values correspond to an additional 12.4% decrease in analgesic use for the average patient treated with LESI and without depression, and a 9.4% decrease for the average treated depressed patient, consistent with the assumptions from Scenario 3. This reduction in the probability of analgesic use among treated patients is in addition to any reduction from the decrease in pain received by treatment. The models for underlying follow-up pain (Model 4) and observed pain (Model 3, with τ_i_ ∼ N(1.5 − 0.5*C*_dep,*i*_, 1.4^2^)) were the same as in Scenario 3.

### Methods used to account for analgesic use

We next used eight candidate analytic methods (A-H) to analyze the simulated datasets and estimate treatment effects on underlying pain (see **Table 1**; further details are provided in Supplemental Table S1). These methods have previously been used in cardiovascular studies examining the effects of an exposure on an outcome in situations where the outcome is affected by concurrent medication use.^13 12-15^. For most methods, one of the following linear regression models was fit on the simulated data:

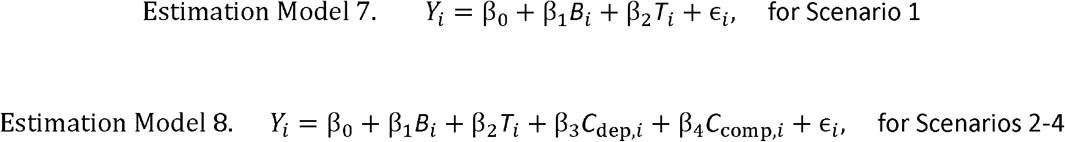

**Table 1.** Candidate analytic methods to account for analgesic use*.

| **Table 1. Candidate analytic methods to account for analgesic use\*** |  |
| --- | --- |
| **Method** | **Description** |
| Method A: Ignoring analgesic use (no adjustment) | Method A assumes that observed pain values are equivalent to underlying pain values. ***This method is used in the vast majority of RCTs of pain treatments, and thus is the "default" comparator method of interest.*** |
| Method B: Adjustment for analgesic use as a covariate | Method B includes an indicator for analgesic use and adjusts for analgesic use as a covariate. |
| Method C: Addition of 1.0 NRS points for analgesic use | Methods C, D, and E compensate for analgesic use by adding a fixed constant to observed pain values, reflecting the improvement in pain NRS produced by analgesic use. This is a form of simple imputation, and can be viewed as a quantitative pain and analgesia composite outcome. The constant of 1.5 was informed by data from the cohort study by Kneeman et al.<sup>21</sup> in which, on average, patients using analgesics reported that their pain when not using analgesics (underlying pain) was ~1.5 NRS points higher than their pain intensity when taking analgesics (observed pain). The addition of constants slightly smaller (1.0) and larger (2.0) than 1.5 was also tested. |
| Method D: Addition of 1.5 NRS points for analgesic use |  |
| Method E: Addition of 2.0 NRS points for analgesic use |  |
| Method F: Exclude analgesic users | Method F excludes patients with concurrent analgesic use at follow-up, analyzing only those who do not use analgesics. |
| Method G: Censored normal regression | Method G uses a parametric survival model and assumes that the observed pain values of patients who use analgesics are right-censored (i.e., analgesic use cannot *increase* pain). |
| Method H: Analgesic use as an outcome: | Method H uses a logistic regression model with analgesic use a binary outcome. Here, analgesic use is a proxy for underlying pain, and the treatment effect is estimated not through reduction of pain but instead through reduction of analgesic use. |
| *Further details are provided in Supplemental Table S1 and the Supplementary Methods |  |

We label these models as “Estimation Models” to distinguish them from the data generation (simulation) models described above (Models 1-6). Note that Estimation Models 7 and 8 are nearly the same as those from which the underlying follow-up pain values *U*_*i*_ were simulated as described above (Model 1 and Model 4), but here the response variable *Y* is based upon the observed pain values *O* instead of the unobserved *U* which represents underlying pain. Each of the methods attempts to handle this discrepancy between the observed and unobserved pain values in a different manner. As a gold standard for comparison, we also fit the ideal model in which we allow *Y*_*i*_ = *U*_*i*_, assuming the underlying pain values were known (which we refer to subsequently as “*knowing underlying pain*”).

### Method A-Ignoring analgesic use (no adjustment for analgesic use)

Method A takes *Y*_*i*_ = *O*_*i*_ for all patients, ignoring all analgesic effects and assuming that the observed pain values are equivalent to the underlying pain values. Method A is the analytic method used in the vast majority of pain RCTs, which conduct ITT analyses without accounting for analgesic use. Therefore, it is the “default” comparator method of interest.

### Method B-Adjustment for analgesic use as a covariate

Method B includes an indicator for analgesic use, modifying the above models (Models 7 and 8) to

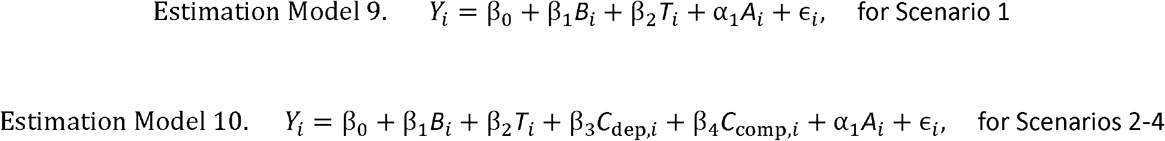

This adjustment is an attempt to tease apart the effect of analgesic use from that of the treatment.

### Methods C, D, and E-Addition of a constant value to analgesic users

Methods C, D, and E compensate for the effect of analgesic use by adding a predetermined constant to the observed pain values, reflecting the expected improvement in pain NRS produced by analgesic use. This is a form of simple imputation and can be viewed as a quantitative pain and analgesia composite outcome; composite outcomes seek to combine impacts on multiple outcomes, and are recognized as alternative primary outcomes in RCTs.^9,10^ Specifically, the response variable is defined by:

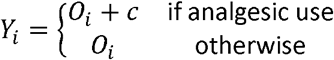

Three different values for the constant, *c* = 1.0, 1.5, and 2.0, were chosen for this study (methods C, D, and E, respectively). The constant of 1.0 was informed empirically, based on meta-analyses of pain RCTs showing treatment effects of analgesics of approximately 1.0 NRS points.^11,12^ The constant of 1.5 was also informed empirically, using data from the above-mentioned study by Kneeman et al.,^4^ in which patients using analgesics reported that their underlying pain intensity was ∼1.5 NRS points higher than their observed pain intensity (unpublished results). In method D, observed pain values were allowed to take half-integers, in contrast to the other simulated data which rounded to the nearest whole number. A larger constant (2.0) was also tested. While the effects of analgesics on pain NRS assumed by methods D and E (1.5 and 2.0 NRS points, respectively) are substantially higher than the mean effects seen in pain RCTs (∼1.0 NRS points, and often smaller),^11,12^ larger mean effects on the pain NRS among analgesic users in actual practice would be expected as compared to mean effects across all participants in an RCT. This is true because in actual practice, individuals who do not have pain relief with initial attempts to use an analgesic will be more likely to stop taking that analgesic. In contrast, those who perceive a benefit of an analgesic in terms of pain relief will be more likely to continue taking that analgesic.

### Method F-Exclude analgesic users

This method simply excludes all patients with indicated analgesic use, performing the regression only upon those patients without analgesic use.

### Method G-Censored normal regression

For this method, a parametric survival model is fit under the assumption that the observed pain values of patients who used analgesics are right-censored (i.e., analgesic use cannot *increase* pain intensity).^5^ That is, we assume that

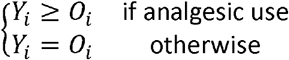

and that the response variable *Y* follows a normal distribution. This model implicitly assumes that the censoring is non-informative, i.e., the mechanism giving rise to the censoring is statistically independent from the underlying uncensored values. Although this assumption is not strictly true in any of these simulation scenarios, censored normal regression is robust to violations of this assumption in other simulation studies.^5^

### Method H-Analgesic use as response

Finally, the following logistic regression model was fit using analgesic use instead of observed pain as a result

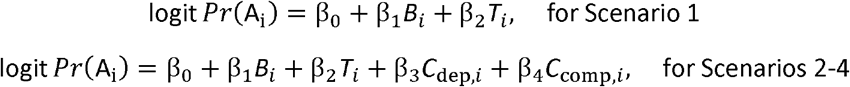

Here analgesic use is used as a proxy for underlying pain, and the treatment effect is estimated not through reduction of pain but instead though reduction of analgesic use.

### Applying methods A-H to the simulated data

For each of the four scenarios, 10,000 simulated data sets were created, and all of the methods (A-H) were applied to each scenario, fitting the models described in the previous section. Additionally, models were fit according to the gold standard model in which underlying pain was assumed to be known. Coefficient estimates were averaged across all simulated data sets, and the standard error for the LESI treatment coefficient obtained by taking the standard deviation of those coefficient estimates. The power to detect a treatment effect of LESI (*β*_2_) was computed by the proportion of 95% confidence intervals for 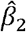 that did not include zero. This is equivalent to rejecting the null hypothesis β_2_ =0 using a two-tailed test at level of significance α= 0.5. To check the type I error for each method, an additional 10,000 data sets were simulated under each scenario with the parameter β_2_ set to zero. Then type I error was estimated in the same manner as power, as the proportion of 95% confidence intervals for 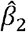 that did not include zero, which should be expected to be near the level of significance α = 0.05 for a valid test. Two additional sets of simulations and model fitting were also conducted as secondary analyses. Whereas the primary simulations assumed that underlying pain and observed pain were the same (correlations of 100%) for participants who were not taking analgesics and truncated NRS values such that no NRS values could be less than 0 or greater than 10, the simulations conducted as part of secondary analysis #1 assumed correlations between underlying pain and observed pain of 95-99% (depending on the simulated dataset) in those not taking analgesics. The simulations conducted as part of secondary analysis #2 also assumed correlations between underlying pain and observed pain of 95-99% in those not taking analgesics, but did not truncate NRS values. Analyses were performed in the R statistical environment (version 4.0.3) and used the package survival version 3.2-7.^13,14^

### Interpretation of the simulation results in terms of power and bias

The simulation results presented in Tables 2 and 3, and Supplemental Tables S2 and S3 (Supplemental File 1), involve two related components: (1) estimation, reflected by the columns for the beta coefficients β_0_ − β_4_, and (2) hypothesis testing, reflected by the “power (%)” and “type I error (%)” columns. Values in these columns for each analytic method can be compared to those yielded by the “ideal” situation in which underlying pain is known. The statistical power of each analytic method (A-H) can be assessed by comparing the value for “power” for the analytic method with that yielded when underlying pain is known. The convention of 80% power is typically used in RCT power calculations and was the default used in the simulations; values lower than 80% have lower power than the typical bar used in RCTs, and values above 80% have greater power.

**Table 2.** (Scenario 1; no covariates): Mean estimates of beta coefficients (expected increase in observed pain intensity NRS outcome per unit increase in predictor variable), power to detect the treatment effect of LESI, and type I error, comparing different methods to account for analgesic use.

| Analysis Method | Coefficient |  |  |  |  | Power (%) | Type I error (%) |
| --- | --- | --- | --- | --- | --- | --- | --- |
| | $\beta_0$<br>Intercept | $\beta_1$<br>Baseline | $\beta_2$<br>LESI (SE) | $\beta_3$<br>Dep. | $\beta_4$<br>Comp. | | |
| Model values (as simulated) | 0.00 | 0.50 | -0.60 |  |  |  |  |
| Knowing underlying pain | 0.35 | 0.46 | -0.56 (0.20) |  |  | 80.0 | 4.9 |
| A. <u>No adjustment*</u> | 0.21 | 0.36 | -0.44 (0.19) |  |  | 64.9 | 4.9 |
| B. Analgesic adjustment as a covariate | 0.16 | 0.35 | -0.43 (0.19) |  |  | 61.9 | 5.0 |
| C. Add 1.0 NRS points | 0.41 | 0.41 | -0.50 (0.20) |  |  | 71.9 | 5.1 |
| D. Add 1.5 NRS points | 0.51 | 0.44 | -0.53 (0.21) |  |  | 72.9 | 5.0 |
| E. Add 2.0 NRS points | 0.62 | 0.46 | -0.56 (0.22) |  |  | 73.2 | 4.9 |
| F. Exclude analgesic | 0.17 | 0.35 | -0.42 (0.25) |  |  | 39.9 | 4.8 |
| G. Censored normal regression | 0.37 | 0.56 | -0.68 (0.28) |  |  | 68.2 | 5.3 |
| H. Analgesic use | -1.24 | 0.21 | -0.25 (0.20) |  |  | 22.6 | 5.3 |

\*the standard analytic method used in pain RCTs.
LESI= lumbar epidural steroid injection, SE = standard error, Dep.= depression, Comp.= seeking disability

**Table 3.** (Scenario 4; treatment affects analgesic use): Mean estimates of beta coefficients (expected increase in observed pain intensity NRS outcome per unit increase in predictor variable), power to detect the treatment effect of LESI, and type I error, comparing different methods to account for analgesic use.

| Analysis Method | Coefficient |  |  |  |  | Power (%) | Type I error (%) |
| --- | --- | --- | --- | --- | --- | --- | --- |
| | $\beta_0$<br>Intercept | $\beta_1$<br>Baseline | $\beta_2$<br>LESI (SE) | $\beta_3$<br>Dep. | $\beta_4$<br>Comp. | | |
| Model values (as simulated) | 0.00 | 0.50 | -0.60 | 0.60 | 0.60 |  |  |
| Knowing underlying pain | 0.32 | 0.46 | -0.56 (0.20) | 0.56 | 0.57 | 79.5 | 4.8 |
| A. No adjustment* | 0.11 | 0.38 | -0.33 (0.19) | 0.87 | 0.47 | 40.4 | 10.4 |
| B. Analgesic adjustment as a covariate | 0.04 | 0.36 | -0.28 (0.19) | 0.91 | 0.45 | 30.6 | 13.1 |
| C. Add 1.0 NRS points | 0.32 | 0.43 | -0.48 (0.20) | 0.74 | 0.52 | 66.5 | 5.3 |
| D. Add 1.5 NRS points | 0.43 | 0.45 | -0.56 (0.21) | 0.67 | 0.55 | 75.8 | 5.1 |
| E. Add 2.0 NRS points | 0.54 | 0.47 | -0.64 (0.22) | 0.60 | 0.58 | 82.4 | 5.9 |
| F. Exclude analgesic | 0.07 | 0.36 | -0.28 (0.24) | 0.76 | 0.45 | 21.0 | 10.0 |
| G. Censored normal | 0.42 | 0.55 | -0.86 (0.27) | 0.41 | 0.70 | 89.9 | 12.8 |
| H. Analgesic use | -1.25 | 0.21 | -0.67 (0.21) | -0.59 | 0.25 | 89.2 | 51.2 |

\*the standard analytic method used in pain RCTs.
LESI= lumbar epidural steroid injection, SE = standard error, Dep.= depression, Comp.= seeking disability

Bias with each analytic method (A-H) can be assessed in two ways. First, this can be done by comparing the estimates of the treatment effect of LESI to the treatment effect for that scenario estimated in the ideal situation where underlying pain is known (the second row in each table). It is important to note that the estimated coefficient values can differ slightly from the true model values (the first row in each table), mostly at the intercept; this is likely due in part to the constraining between 0 and 10 and rounding to the nearest whole number of pain NRS values. As an example to guide interpretation, if the estimated effect of LESI is -0.56 (reflecting the between-group difference in pain intensity with LESI vs. lidocaine-alone epidural injections in the model when underlying pain is known in the LESS RCT), values between -0.56 and 0 indicate some degree of bias towards a null effect, and values that are more negative than -0.56 indicate bias in the opposite direction. Second, information about bias- as reflected by invalid statistical inference- is also contained in the values for “type I error” generated with each analytic method; values for type I error larger than the commonly used significance level of 5% indicate that hypothesis testing will incorrectly reject a true null hypothesis more than 5% of the time with the analytic method, i.e., finding a statistically significant effect of treatment where no such effect exists. Methods can result in biased estimates for the treatment effect of LESI while still maintaining valid inference when estimates are shrunk toward zero under alternative hypotheses, but remain unbiased under the null hypothesis. An analytic method will be useful if it optimizes power (as close to 80% as possible, or higher), while still being unbiased (beta coefficients are similar to those from the model where underlying pain is known, type I error is ∼5%). A method with adequate (or more than adequate) power is not useful if it rejects the hypothesis too often under the null hypothesis (i.e., if type I error is >5%).

### Analyses of real (non-simulated) RCT data

We next analyzed individual-level patient data from the LESS RCT. The LESS study design, measures, and analytic methods have been reported previously.^18^ Post-randomization opioid use was ascertained using patient diaries completed daily for the first 3 weeks after epidural injection in a subset of the LESS sample (n=335). LESS participants who reported opioid use at or in the 3 days prior to the 3-week post-randomization assessment were classified as having “any analgesic use,” while those who had no opioid use in this period were classified as having “no analgesic use”; non-opioid analgesic use data was not available and therefore not part of the definition of any analgesic use at 3 weeks post-randomization. Analytic methods A-H were then applied to examine associations between the LESI treatment and leg pain intensity at 3-week follow-up. Regression analyses were conducted comparing leg pain intensity NRS scores at 3-weeks post randomization according to randomized group (LESI vs control epidural injections of lidocaine alone), adjusting for study site and baseline leg pain NRS, using linear regression for Methods A-F, censored normal regression (Tobit regression) for Method G, and logistic regression for Method H. Each method was evaluated by examining the statistical significance of associations between the LESI treatment and leg pain intensity at 3-week follow-up, with lower p-values interpreted as indicators of greater power. SAS version 9.4 (Cary, North Carolina) was used for these analyses.

## RESULTS

### Scenario 1 (no covariates, Table 2)

The treatment effect of LESI under the gold standard when underlying pain is known was -0.56 (improvement in pain with LESI), representing a slight decrease from the true model value of -0.6 NRS points. When knowing underlying pain, 80% power was attained; as expected, all analytic methods (A-H) fell short of that achieved when knowing underlying pain. The default analysis method used in studies of pain, ignoring analgesic use (method A), resulted in “shrinkage” of the estimated effect of LESI, with a regression coefficient (−0.44) that was smaller in magnitude than the value from the model when underlying pain was known (−0.56), resulting in 65% power. Adjustment for analgesic use as a covariate (Method B) also resulted in decreased power (62%). All other methods (C-H) also yielded power that was less than 80%, and in most of these cases, the LESI treatment coefficient estimates were shrunk relative to the model value when underlying pain was known (−0.56). Power was lowest with analyzing analgesic use as a binary outcome (method H, 23% power). The methods with highest power were those that added 1.5 or 2.0 NRS points to observed pain values for those with concurrent analgesic use (methods D and E), both of which had 73% power. These two methods also resulted in minimal to no shrinkage of the estimated LESI effect (−0.53 and -0.56, respectively), as compared to the model value when underlying pain was known (−0.56). Type I error was near 5% for all methods, indicating valid inference.

### Scenario 2 (covariates associated with underlying pain, Supplemental Table S2)

Adding the two baseline covariates for depression and receiving disability compensation produced very similar results to those from Scenario 1 (Supplemental Table S2). Under the gold standard of knowing underlying pain, power was 80%. Method A (ignoring analgesic use) had 64% power, and method B (adjustment for analgesic use as a covariate) had 61% power. The methods with highest power were again methods D and E (adding 1.5 or 2.0 NRS points, respectively, to observed pain values when analgesic use was present), both which had 72% power. These two methods also resulted in minimal to no shrinkage of the estimated LESI effect (−0.54 and -0.56, respectively, compared to -0.56 when underlying pain was known). Type I error was near 5% for all methods, indicating valid inference.

### Scenario 3 (covariate for analgesic use, Supplemental Table S3)

Allowing the depression covariate to affect probability of analgesic use and size of analgesic effect also produced very similar results to those from Scenario 1 and 2 (Supplemental Table S3). Method A and B power of 65% and 62%, respectively, and the methods with highest power were again methods D and E, both of which had 73% power with minimal to no shrinkage of the estimated LESI effect (−0.54 and -0.57, respectively, compared to -0.56 when underlying pain was known). Type I error was near 5% for all methods.

### Scenario 4 (treatment affects analgesic use, Table 3)

Allowing the LESI treatment variable to affect the probability of analgesic use changed the simulation study results dramatically, as compared to Scenarios 1-3. Most notably, for nearly every method except Methods C, D, and E, type I error was substantially inflated above 5%, so inference was no longer valid (Table 3). Because of this, power values with the different analytic methods in Scenario 4 should be ignored. Adding the “correct” 1.5 NRS constant to observed pain (method D) provided approximately valid inference in Scenario 4 (5% type I error), and there were very small increases in type I error when this constant was misspecified as 1.0 or 2.0. Thus, in Scenario 4, the method of adding a constant value to the NRS of patients with concurrent analgesic use works best when the constant precisely estimates the mean analgesic effect.

The results from secondary analyses #1 and #2, examining different assumptions regarding the truncation of NRS values and correlations between underlying pain and observed pain, are presented in Supplemental File 2 (Supplemental Table S4-S11). These results showed no material differences from the primary analyses.

### Analysis of real data from LESS

In analyses of data from the LESS RCT, the subgroups of those randomized to LESI vs. epidural injections of lidocaine alone were comparable with regards to sociodemographic and clinical characteristics (Supplemental Table S12). Ignoring analgesic use (Method A), produced an estimated treatment effect of LESI of -0.58 NRS points, and a p-value of 0.03 (Table 4). With adjustment for analgesic use as a covariate (Method B), there was shrinkage of the estimated LESI effect (−0.42 NRS points) and less apparent power (*p* = 0.12), relative to Method A, and a similar pattern was also seen for Method F (excluding analgesic users). The estimated treatment effects of LESI were larger and statistical significance was greater for Methods C, D and E, relative to Method A, including a LESI effect of -0.80 NRS points with Method D (*p* = 0.007), which adds 1.5 NRS points to observed pain intensity among those with concurrent analgesic use. The statistical significance of results using Method G was generally similar to that produced by Methods C-E, and the statistical significance of results using Method H was similar to that produced with Method A (Table 4).

**Table 4.** Analysis of real (non-simulated) data from the LESS RCT.

| **Analysis Method** | **Treatment effect of LESI**<br>**(95% CI)** | **p-value** |
| --- | --- | --- |
| A. Ignoring analgesic use* | -0.58 (-1.12, 0.04) | 0.03 |
| B. Analgesic adjustment<br>as a covariate | -0.42 (-0.96, 0.11) | 0.12 |
| C. Add 1.0 NRS points | -0.73 (-1.29, -0.17) | 0.01 |
| D. Add 1.5 NRS points | -0.80 (-1.38, -0.22) | 0.007 |
| E. Add 2.0 NRS points | -0.88 (-1.48, -0.28) | 0.004 |
| F. Exclude analgesic<br>users | -0.47 (-1.12, 0.19) | 0.16 |
| G. Censored normal<br>regression | -0.71 (-1.20, -0.23) | 0.004 |
| H. Analgesic use as a<br>binary outcome | 0.56 (-0.35-0.89)<sup>a</sup> | 0.01 |

Negative values for treatment effects reflect benefit with LESI (versus epidural injections of lidocaine) on NRS pain at 3-week follow-up, with smaller p-values interpreted as greater apparent power to detect the treatment effect of LESI with the analysis method. Analgesic use reflects opioid use only; data on non-opioid analgesic use was not available.
LESS RCT= Lumbar Epidural Steroid Injections for spinal Stenosis randomized controlled trial, LESI= lumbar epidural steroid injection, CI= confidence interval \*the standard analytic method used in pain RCTs.
<sup>a</sup>Odds ratio for the effect of LESI on opioid use as a binary outcome, reflecting less opioid use in those randomized to LESI

## DISCUSSION

In this study, we compared analytic methods to account for analgesic use in pain RCTs, first in analyses of simulated RCT data (which can inform about both power and bias), and second in analyses of real RCTs (which can inform about power, but not bias).^17^ In simulations, compared to the gold standard when underlying pain intensity was known (power ∼80%), power to detect a treatment effect of LESI was decreased with all analytic methods studied (power ∼22% to 73%). Most methods also demonstrated “shrinkage” of the LESI treatment effect, causing it to appear smaller than it actually was. Compared to the ideal situation when underlying pain intensity is known, power was substantially lower with Method A (ignoring analgesic use: 63% to 65% power in Scenarios 1-3), the default analytic method used in pain RCTs. Power was also lower with Method B (adjusting for analgesic use as a covariate: power 61% to 62% in Scenarios 1-3), and markedly lower with Method H (analyzing analgesic use as a binary outcome: power ∼22%). Of the methods examined, Methods C, D, and E (addition of an empirically justified fixed constant to the observed pain NRS among those with concurrent analgesic use) best optimized power (power 71% to 73% for Scenarios 1-3). Bias was negligible with all methods examined in Scenarios 1-3, however, in the more complex setting of Scenario 4, most of the analytic methods led to invalid inference, and only Method D (addition of 1.5 to the observed pain NRS among those with concurrent analgesic use) had negligible bias. The utility of Method D was further supported by analysis of real data from the LESS RCT, in which Method D had greater statistical significance than most other methods examined.

Further consideration of the relationships modeled in our simulations can help to illustrate how the common practice of ignoring analgesic use in conventional ITT analyses of pain intensity estimates the “total effect” of a treatment, which is biased towards the null, and is distinct from the “controlled direct effect,” which may be of greater interest to clinicians and patients (Figure 1). All simulations assumed two different ways in which the LESI treatment may affect observed pain: (1) via direct effects on pain, and (2) via indirect effects on pain mediated by changes in analgesic use (Figure 1). In this context, because LESI lowers pain on average for those receiving it, those receiving the control treatment have greater pain and are therefore more likely to use concurrent analgesics, with consequent improvements in pain due to analgesic use. Thus, when ignoring analgesic use (Method A), the *observed* pain intensity values for participants in the control group appear lower than they should be when compared to the corresponding observed pain values in those receiving LESI; analysis of these *observed* pain intensity values estimates the *total effect* of LESI. In contrast, Method D calculates *underlying pain* intensity values by adding 1.5 to the observed pain NRS among those with concurrent analgesic use, and assumes that underlying and observed pain are equal among those not using analgesics; analysis of these *underlying* pain intensity values estimates the *controlled direct effect* of LESI. Therefore, while our simulations and analyses of real data indicate attenuation of the LESI effect on pain intensity with Method A relative to Method D, the two methods actually reflect two different types of effects (estimands). In the context of an explanatory (efficacy) trial where all participants are strictly monitored to prevent any use of analgesics other than the randomized treatment, the total effect and controlled direct effect may be equivalent. In the context of a pragmatic trial, however, the total effect and controlled direct effect are likely to differ. Importantly, adjustment for analgesic use as a covariate (Method B) in the current study did *not* improve upon ignoring analgesic use in analyses of simulated or real RCT data, and such adjustment further attenuated effect sizes. This may be a consequence of inappropriate adjustment for a mediator, a known epidemiological phenomenon which can induce bias in some settings,^25^ and illustrates that finding alternatives to the status quo of ignoring analgesic use is not necessarily a simple matter.

We propose that imputation of underlying pain with the addition of an appropriate fixed constant to observed pain among those with concurrent analgesic use should be considered as an alternative analytic approach to ignoring analgesic use in pain RCTs. The addition of 1.5 points to observed pain produces a **q**uantitative **p**ain and **a**nalgesic **c**omposite outcome, which for simplicity we will refer to as the QPAC_1.5_. Composite outcomes have shown promise as an alternate outcome measure in RCTs of treatments for neuropathic pain.^26^ However, many composite outcomes are binary outcomes, which can limit statistical power. While a composite outcome, the QPAC_1.5_ preserves the NRS as a quasi-continuous variable, permitting greater statistical power while making explicit the assumption that the average effect of concurrent analgesic use on observed pain among analgesic users is 1.5 NRS points. The QPAC_1.5_ can be retrospectively applied to any existing RCT dataset in which the NRS and concurrent analgesic use were evaluated contemporaneously. Our experience as pain trialists has been that such information is already collected in many pain RCTs, yet is rarely fully utilized. Another potential advantage of the QPAC_1.5_ is that it can easily be modified if clinical knowledge or research indicates that an alternative to 1.5 is more appropriate in a particular population. Although our simulations suggested relatively small effects on bias with misspecification of the fixed constant that proxies the effect of concurrent analgesics (e.g. Methods C and E in Tables 2 and 3), further research is warranted to determine the optimal constant in different settings.

A strength of the QPAC_1.5_ is that it can be easily used without specialized statistical expertise. On the other hand, the QPAC_1.5_ is an oversimplification, since it assumes the same average effect of analgesics among all individuals. Future research may use simple imputation methods (like that used to calculate the QPAC_1.5_) to improve upon the QPAC_1.5_ by accounting for the use of different analgesic classes, doses, or multiple analgesics. Other contemporary methods may have even greater utility. Given the success of simple imputation with the QPAC_1.5_, multiple imputation should be considered.^13^ Inverse probability weighting or “g-methods” may also be used to account for concurrent treatments,^27,28^ and have not been widely used in pain research. However, these contemporary methods will require greater knowledge about predictors of actual analgesic *use* than is currently available (as most published studies of analgesics analyze pharmacy data, which are informative only about what is *prescribed*^8^). The constantly changing nature of pain and the ability of patients to self-regulate analgesic use without physician visits or prescriptions also may require frequent, repeated reassessment of variables, posing data collection challenges. While the QPAC_1.5_ is not intended to be the definitive solution to the problem of how to account for analgesic use in pain RCTs, it represents an improvement over the current commonly used methods and it can be further refined in future work.

A limitation of the current study is that the findings may apply only to situations where treatment effects and analgesic effects are modest and of generally comparable magnitude. They may not apply to contexts where the pain treatment under study has a large effect on pain (e.g., knee replacement), or where analgesic effects are very large (e.g., post-surgical pain management with parenteral medications). On the other hand, although the current study simulated a specific treatment for a specific condition (LESI for LSS), the relationships modeled (see Supplemental Methods) are common to most pain treatments and conditions, and are by no means specific to the situation of LESI. Therefore, we expect the findings to be broadly applicable to many situations where pain treatments are delivered on an outpatient basis.

In this study, we used a contemporary causal inference framework to formalize alternative estimation goals (estimands) that incorporate analgesic use into the primary outcomes of pain treatment RCTs, and examined analytic methods that can potentially account for concurrent analgesic use. In analyses of simulated and real RCT data, we found that power to detect an effect of treatment with LESI was decreased with all commonly used analytic methods of accounting for concurrent analgesic use, relative to the ideal situation where patients’ underlying pain intensity was known for all participants. This loss of power was minimized by using a simple quantitative pain and analgesia composite outcome, which we have named the QPAC_1.5_. The QPAC_1.5_ may have promise for optimizing power to detect the effects of pain treatments in situations where concurrent analgesic treatment is expected, and should be considered as an alternative analytic method to be used in pain treatment RCTs.

## Data Availability

All data produced in the present study are available upon reasonable request to the authors.

