## Supplemental Tables for "Improving Power and Accuracy in Randomized Controlled Trials of Pain Treatments by Accounting for Concurrent Analgesic Use"

| Supplemental Table S1. Detailed descriptions of candidate analytic methods to account for analgesic use, including models | |
|---|---|
| **Method** | **Description** |
| Method A: Ignoring analgesic use (no adjustment) | Method A takes $Y_i = O_i$ for all patients, ignoring all analgesic effects and assuming that the observed pain values are equivalent to the underlying pain values. ***Method A is used in the vast majority of RCTs of pain treatments, and thus is the "default" comparator method of interest.*** |
| Method B: Adjustment for analgesic use as a covariate | Method B includes an indicator for analgesic use, such as $Y_i = \beta_0 + \beta_1 B_i + \beta_2 T_i + \alpha_1 A_i + \epsilon_i$(Scenario 1), and $Y_i = \beta_0 + \beta_1 B_i + \beta_2 T_i + \beta_3 C_{\mathrm{dep},i} + \beta_4 C_{\mathrm{comp},i} + \alpha_1 A_i + \epsilon_i$(Scenario 2-4) |
| Method C, D, and E: Addition of a fixed constant value (1.0, 1.5, and 2.0, respectively) for analgesic users | Method C, D, and E compensate for analgesics use by adding a fixed constant to observed pain values, reflecting the improvement in pain NRS produced by analgesic use. This is a form of simple imputation, and can be viewed as a quantitative pain and analgesia composite outcome. Specifically, the response variable is defined by: $$Y_i = \begin{cases} O_i + c & \text{if analgesic use} \\ O_i & \text{otherwise} \end{cases}$$ Three different values for the constant, *c* = 1.0, 1.5, and 2.0, were used (methods C, D, and E, respectively). The constant of 1.5 was informed by data from a cohort study conducted by our group (Kneeman et al.)[1] in which patients using analgesics reported that their pain intensity when not using analgesics (underlying pain) was ~1.5 NRS points higher than their pain intensity when taking analgesics (observed pain); the addition of constants slightly smaller (1.0) and larger (2.0) than 1.5 was also tested. |
| Method F: Exclude analgesic users | Method F excludes all patients with indicated analgesic use, performing the regression only upon those patients without concurrent analgesic use at follow-up. |
| Method G: Censored normal regression | In method G, a parametric survival model is fit assuming that the observed pain values of patients who used analgesics are right-censored (i.e., analgesic use cannot *increase* pain intensity). That is: $$\begin{cases} Y_i \geq O_i & \text{if analgesic use} \\ Y_i = O_i & \text{otherwise} \end{cases}$$ The response variable *Y* follows a normal distribution. This model implicitly assumes that censoring is non-informative, however censored normal regression is robust to violations of this assumption.[2] |
| Method H: Analgesic use as an outcome: | In method H, a logistic regression model is fit using analgesic use the outcome (instead of observed pain): $\text{logit}\, Pr(\mathrm{A_i}) = \beta_0 + \beta_1 B_i + \beta_2 T_i$ (Scenario 1), and $\text{logit}\, Pr(\mathrm{A_i}) = \beta_0 + \beta_1 B_i + \beta_2 T_i + \beta_3 C_{\mathrm{dep},i} + \beta_4 C_{\mathrm{comp},i}$ (Scenarios 2-4). Here analgesic use is used as a proxy for underlying pain, and the treatment effect is estimated not through reduction of pain but instead though reduction of analgesic use. |

**Supplemental Table S2 (Scenario 2; also including covariates associated with underlying pain):** Mean estimates of beta coefficients (expected increase in observed pain intensity NRS outcome per unit increase in predictor variable), power to detect the treatment effect of LESI, and type I error, comparing different methods to account for analgesic use.

| Analysis Method | $\beta_0$ Intercept | $\beta_1$ Baseline | $\beta_2$ LESI (SE) | $\beta_3$ Dep. | $\beta_4$ Comp. | Power (%) | Type I error (%) |
|---|---|---|---|---|---|---|---|
| | Coefficient | | | | | | |
| Model values (as simulated) | 0.00 | 0.50 | -0.60 | 0.60 | 0.60 | | |
| Knowing underlying pain | 0.32 | 0.46 | -0.56 (0.20) | 0.56 | 0.57 | 79.5 | 4.8 |
| A. Ignoring analgesic use* | 0.17 | 0.37 | -0.45 (0.19) | 0.45 | 0.46 | 64.3 | 5.1 |
| B. Analgesic adjustment as a covariate | 0.12 | 0.36 | -0.43 (0.19) | 0.44 | 0.44 | 61.4 | 5.0 |
| C. Add 1.0 NRS points | 0.38 | 0.42 | -0.51 (0.20) | 0.51 | 0.52 | 71.0 | 4.9 |
| D. Add 1.5 NRS points | 0.48 | 0.44 | -0.54 (0.21) | 0.54 | 0.55 | 72.1 | 5.0 |
| E. Add 2.0 NRS points | 0.58 | 0.47 | -0.56 (0.22) | 0.57 | 0.57 | 72.2 | 5.0 |
| F. Exclude analgesic users | 0.14 | 0.35 | -0.43 (0.26) | 0.44 | 0.44 | 39.0 | 5.0 |
| G. Censored normal regression | 0.30 | 0.57 | -0.69 (0.29) | 0.72 | 0.73 | 67.1 | 5.0 |
| H. Analgesic use at follow-up (binary outcome) | -1.25 | 0.21 | -0.25 (0.21) | 0.25 | 0.26 | 22.1 | 5.0 |

*the standard analytic method used in pain RCTs. LESI= lumbar epidural steroid injection, SE = standard error, Dep.= depression, Comp.= seeking disability

**Supplemental Table S3 (Scenario 3; also including covariates associated with analgesic use):** Mean estimates of beta coefficients (expected increase in observed pain intensity NRS outcome per unit increase in predictor variable), power to detect the treatment effect of LESI, and type I error, comparing different methods to account for analgesic use.

| Analysis Method | $\beta_0$ Intercept | $\beta_1$ Baseline | $\beta_2$ LESI (SE) | $\beta_3$ Dep. | $\beta_4$ Comp. | Power (%) | Type I error (%) |
|---|---|---|---|---|---|---|---|
| | Coefficient | | | | | | |
| Model values (as simulated) | 0.00 | 0.50 | -0.60 | 0.60 | 0.60 | | |
| Knowing underlying pain | 0.35 | 0.46 | -0.56 (0.20) | 0.56 | 0.57 | 79.5 | 4.8 |
| A. Ignoring analgesic use* | 0.14 | 0.37 | -0.45 (0.19) | 0.88 | 0.46 | 65.4 | 5.0 |
| B. Analgesic adjustment as a covariate | 0.07 | 0.36 | -0.43 (0.19) | 0.93 | 0.45 | 61.9 | 5.0 |
| C. Add 1.0 NRS points | 0.34 | 0.42 | -0.51 (0.20) | 0.75 | 0.52 | 71.6 | 4.9 |
| D. Add 1.5 NRS points | 0.45 | 0.45 | -0.54 (0.21) | 0.68 | 0.55 | 72.7 | 4.7 |
| E. Add 2.0 NRS points | 0.55 | 0.47 | -0.57 (0.22) | 0.61 | 0.58 | 72.7 | 4.8 |
| F. Exclude analgesic users | 0.12 | 0.36 | -0.43 (0.25) | 0.77 | 0.45 | 40.5 | 5.1 |
| G. Censored normal regression | 0.34 | 0.56 | -0.68 (0.28) | 0.37 | 0.72 | 68.6 | 5.0 |
| H. Analgesic use at follow-up (binary outcome) | -1.25 | 0.21 | -0.25 (0.21) | -0.58 | 0.25 | 22.4 | 4.9 |

*the standard analytic method used in pain RCTs. LESI= lumbar epidural steroid injection, SE = standard error, Dep.= depression, Comp.= seeking disability

**Supplemental Table S4 (Scenario 1; no covariates; no constraining or rounding):** Mean estimates of beta coefficients (expected increase in observed pain intensity NRS outcome per unit increase in predictor variable), power to detect the treatment effect of LESI, and type I error, comparing different methods to account for analgesic use. Pain values were not constrained to between 0 and 10 or rounded to the nearest whole number for these simulations.

| | Coefficient | | | | | | |
|---|---|---|---|---|---|---|---|
| **Analysis Method** | ***β₀*** **Intercept** | ***β₁*** **Baseline** | ***β₂*** **LESI (SE)** | ***β₃*** **Dep.** | ***β₄*** **Comp.** | **Power (%)** | **Type I error (%)** |
| Model values (as simulated) | 0.00 | 0.50 | -0.60 | | | | |
| Knowing underlying pain | 0.00 | 0.50 | -0.60 (0.21) | | | 81.7 | 4.8 |
| A. Ignoring analgesic use* | -0.33 | 0.42 | -0.51 (0.21) | | | 66.9 | 5.0 |
| B. Analgesic adjustment as a covariate | -0.37 | 0.41 | -0.49 (0.21) | | | 64.7 | 5.1 |
| C. Add 1.0 NRS points | -0.12 | 0.47 | -0.57 (0.22) | | | 73.2 | 5.0 |
| D. Add 1.5 NRS points | -0.02 | 0.49 | -0.60 (0.23) | | | 74.5 | 5.1 |
| E. Add 2.0 NRS points | 0.09 | 0.52 | -0.62 (0.24) | | | 74.8 | 5.0 |
| F. Exclude analgesic users | -0.36 | 0.41 | -0.49 (0.28) | | | 43.2 | 4.7 |
| G. Censored normal regression | -0.10 | 0.63 | -0.76 (0.31) | | | 70.2 | 5.0 |
| H. Analgesic use at follow-up (binary outcome) | -1.24 | 0.21 | -0.25 (0.21) | | | 22.8 | 5.1 |

*the standard analytic method used in pain RCTs. LESI= lumbar epidural steroid injection, SE = standard error, Dep.= depression, Comp.= seeking disability

**Supplemental Table S5 (Scenario 2; also including covariates associated with underlying pain; no constraining or rounding):** Mean estimates of beta coefficients (expected increase in observed pain intensity NRS outcome per unit increase in predictor variable), power to detect the treatment effect of LESI, and type I error, comparing different methods to account for analgesic use. Pain values were not constrained to between 0 and 10 or rounded to the nearest whole number for these simulations.

| | Coefficient | | | | | | |
|---|---|---|---|---|---|---|---|
| **Analysis Method** | ***β₀*** **Intercept** | ***β₁*** **Baseline** | ***β₂*** **LESI (SE)** | ***β₃*** **Dep.** | ***β₄*** **Comp.** | **Power (%)** | **Type I error (%)** |
| Model values (as simulated) | 0.00 | 0.50 | -0.60 | 0.60 | 0.60 | | |
| Knowing underlying pain | 0.00 | 0.50 | -0.60 (0.21) | 0.60 | 0.60 | 81.3 | 5.0 |
| A. Ignoring analgesic use* | -0.33 | 0.42 | -0.51 (0.21) | 0.50 | 0.51 | 66.4 | 5.1 |
| B. Analgesic adjustment as a covariate | -0.38 | 0.41 | -0.49 (0.21) | 0.49 | 0.50 | 64.0 | 5.1 |
| C. Add 1.0 NRS points | -0.13 | 0.47 | -0.56 (0.22) | 0.56 | 0.57 | 72.8 | 5.0 |
| D. Add 1.5 NRS points | -0.02 | 0.50 | -0.59 (0.23) | 0.59 | 0.60 | 74.3 | 4.9 |
| E. Add 2.0 NRS points | 0.08 | 0.52 | -0.62 (0.24) | 0.62 | 0.63 | 74.6 | 4.8 |
| F. Exclude analgesic users | -0.36 | 0.41 | -0.49 (0.28) | 0.49 | 0.49 | 41.1 | 5.0 |
| G. Censored normal regression | -0.14 | 0.64 | -0.76 (0.31) | 0.78 | 0.80 | 69.3 | 5.1 |
| H. Analgesic use at follow-up (binary outcome) | -1.25 | 0.21 | -0.25 (0.21) | 0.25 | 0.26 | 22.7 | 4.6 |

*the standard analytic method used in pain RCTs. LESI= lumbar epidural steroid injection, SE = standard error, Dep.= depression, Comp.= seeking disability

**Supplemental Table S6 (Scenario 3; also including covariates associated with analgesic use; no constraining or rounding):** Mean estimates of beta coefficients (expected increase in observed pain intensity NRS outcome per unit increase in predictor variable), power to detect the treatment effect of LESI, and type I error, comparing different methods to account for analgesic use. Pain values were not constrained to between 0 and 10 or rounded to the nearest whole number for these simulations.

| | Coefficient | | | | | | |
|---|---|---|---|---|---|---|---|
| **Analysis Method** | ***β₀*** **Intercept** | ***β₁*** **Baseline** | ***β₂*** **LESI (SE)** | ***β₃*** **Dep.** | ***β₄*** **Comp.** | **Power (%)** | **Type I error (%)** |
| Model values (as simulated) | 0.00 | 0.50 | -0.60 | 0.60 | 0.60 | | |
| Knowing underlying pain | 0.00 | 0.50 | -0.60 (0.21) | 0.60 | 0.60 | 81.3 | 5.0 |
| A. Ignoring analgesic use* | -0.35 | 0.43 | -0.51 (0.21) | 0.98 | 0.51 | 67.1 | 5.0 |
| B. Analgesic adjustment as a covariate | -0.41 | 0.41 | -0.49 (0.21) | 1.01 | 0.50 | 64.5 | 5.1 |
| C. Add 1.0 NRS points | -0.15 | 0.47 | -0.57 (0.22) | 0.84 | 0.57 | 73.3 | 4.8 |
| D. Add 1.5 NRS points | -0.04 | 0.50 | -0.60 (0.23) | 0.77 | 0.60 | 74.8 | 4.7 |
| E. Add 2.0 NRS points | 0.06 | 0.52 | -0.63 (0.24) | 0.71 | 0.63 | 75.0 | 4.7 |
| F. Exclude analgesic users | -0.37 | 0.41 | -0.49 (0.27) | 0.84 | 0.50 | 43.3 | 5.2 |
| G. Censored normal regression | -0.09 | 0.63 | -0.75 (0.30) | 0.40 | 0.78 | 70.4 | 5.1 |
| H. Analgesic use at follow-up (binary outcome) | -1.25 | 0.21 | -0.25 (0.21) | -0.58 | 0.25 | 22.3 | 4.8 |

*the standard analytic method used in pain RCTs. LESI= lumbar epidural steroid injection, SE = standard error, Dep.= depression, Comp.= seeking disability

**Supplemental Table S7 (Scenario 4; treatment affects analgesic use; no constraining or rounding):** Mean estimates of beta coefficients (expected increase in observed pain intensity NRS outcome per unit increase in predictor variable), power to detect the treatment effect of LESI, and type I error, comparing different methods to account for analgesic use. Pain values were not constrained to between 0 and 10 or rounded to the nearest whole number for these simulations.

| | Coefficient | | | | | | |
|---|---|---|---|---|---|---|---|
| **Analysis Method** | ***β₀*** **Intercept** | ***β₁*** **Baseline** | ***β₂*** **LESI (SE)** | ***β₃*** **Dep.** | ***β₄*** **Comp.** | **Power (%)** | **Type I error (%)** |
| Model values (as simulated) | 0.00 | 0.50 | -0.60 | 0.60 | 0.60 | | |
| Knowing underlying pain | 0.00 | 0.50 | -0.60 (0.21) | 0.60 | 0.60 | 81.3 | 5.0 |
| A. Ignoring analgesic use* | -0.36 | 0.43 | -0.36 (0.21) | 0.95 | 0.51 | 40.6 | 10.9 |
| B. Analgesic adjustment as a covariate | -0.42 | 0.41 | -0.32 (0.21) | 0.99 | 0.50 | 32.8 | 13.2 |
| C. Add 1.0 NRS points | -0.15 | 0.47 | -0.52 (0.22) | 0.82 | 0.57 | 65.6 | 5.8 |
| D. Add 1.5 NRS points | -0.04 | 0.50 | -0.59 (0.23) | 0.75 | 0.60 | 74.8 | 5.0 |
| E. Add 2.0 NRS points | 0.07 | 0.52 | -0.67 (0.24) | 0.69 | 0.63 | 81.1 | 5.3 |
| F. Exclude analgesic users | -0.40 | 0.42 | -0.32 (0.27) | 0.83 | 0.50 | 23.1 | 9.9 |
| G. Censored normal regression | 0.01 | 0.60 | -0.94 (0.29) | 0.44 | 0.75 | 90.9 | 12.4 |
| H. Analgesic use at follow-up (binary outcome) | -1.25 | 0.21 | -0.67 (0.21) | -0.59 | 0.25 | 88.9 | 51.3 |

*the standard analytic method used in pain RCTs. LESI= lumbar epidural steroid injection, SE = standard error, Dep.= depression, Comp.= seeking disability

**Supplemental Table S8 (Scenario 1; no covariates; $O_i \neq U_i$):** Mean estimates of beta coefficients (expected increase in observed pain intensity NRS outcome per unit increase in predictor variable), power to detect the treatment effect of LESI, and type I error, comparing different methods to account for analgesic use. Instead of having observed $O_i$ equal unobserved $U_i$ exactly for all patients $i = 1, \dots, n$ without analgesic use, $U_i$ was simulated to have 95-99% correlation with $O_i$ for these simulations.

| | Coefficient | | | | | | |
|---|---|---|---|---|---|---|---|
| **Analysis Method** | ***β₀*** **Intercept** | ***β₁*** **Baseline** | ***β₂*** **LESI (SE)** | ***β₃*** **Dep.** | ***β₄*** **Comp.** | **Power (%)** | **Type I error (%)** |
| Model values (as simulated) | 0.00 | 0.50 | -0.60 | | | | |
| Knowing underlying pain | 0.35 | 0.46 | -0.55 (0.20) | | | 79.5 | 5.1 |
| A. Ignoring analgesic use* | 0.22 | 0.36 | -0.44 (0.19) | | | 63.3 | 5.2 |
| B. Analgesic adjustment as a covariate | 0.17 | 0.35 | -0.42 (0.19) | | | 60.4 | 5.2 |
| C. Add 1.0 NRS points | 0.42 | 0.41 | -0.50 (0.20) | | | 69.8 | 5.1 |
| D. Add 1.5 NRS points | 0.52 | 0.44 | -0.53 (0.21) | | | 71.0 | 5.0 |
| E. Add 2.0 NRS points | 0.62 | 0.46 | -0.56 (0.22) | | | 71.1 | 4.9 |
| F. Exclude analgesic users | 0.19 | 0.35 | -0.42 (0.25) | | | 38.7 | 5.1 |
| G. Censored normal regression | 0.38 | 0.56 | -0.68 (0.28) | | | 66.7 | 4.9 |
| H. Analgesic use at follow-up (binary outcome) | -1.24 | 0.21 | -0.25 (0.20) | | | 22.2 | 4.9 |

*the standard analytic method used in pain RCTs. LESI= lumbar epidural steroid injection, SE = standard error, Dep.= depression, Comp.= seeking disability

**Supplemental Table S9 (Scenario 2; also including covariates associated with underlying pain; $O_i \neq U_i$):** Mean estimates of beta coefficients (expected increase in observed pain intensity NRS outcome per unit increase in predictor variable), power to detect the treatment effect of LESI, and type I error, comparing different methods to account for analgesic use. Instead of having observed $O_i$ equal unobserved $U_i$ exactly for all patients $i = 1, \dots, n$ without analgesic use, $U_i$ was simulated to have 95-99% correlation with $O_i$ for these simulations.

| | Coefficient | | | | | | |
|---|---|---|---|---|---|---|---|
| **Analysis Method** | ***β₀*** **Intercept** | ***β₁*** **Baseline** | ***β₂*** **LESI (SE)** | ***β₃*** **Dep.** | ***β₄*** **Comp.** | **Power (%)** | **Type I error (%)** |
| Model values (as simulated) | 0.00 | 0.50 | -0.60 | 0.60 | 0.60 | | |
| Knowing underlying pain | 0.31 | 0.46 | -0.56 (0.20) | 0.57 | 0.56 | 78.7 | 5.2 |
| A. Ignoring analgesic use* | 0.18 | 0.37 | -0.45 (0.20) | 0.46 | 0.45 | 63.6 | 5.1 |
| B. Analgesic adjustment as a covariate | 0.13 | 0.36 | -0.43 (0.20) | 0.44 | 0.44 | 60.4 | 5.1 |
| C. Add 1.0 NRS points | 0.38 | 0.42 | -0.51 (0.20) | 0.52 | 0.51 | 70.2 | 5.2 |
| D. Add 1.5 NRS points | 0.49 | 0.44 | -0.53 (0.21) | 0.55 | 0.54 | 71.3 | 5.3 |
| E. Add 2.0 NRS points | 0.59 | 0.47 | -0.56 (0.22) | 0.58 | 0.57 | 71.7 | 5.3 |
| F. Exclude analgesic users | 0.16 | 0.35 | -0.42 (0.26) | 0.44 | 0.43 | 36.8 | 4.9 |
| G. Censored normal regression | 0.31 | 0.57 | -0.70 (0.29) | 0.73 | 0.72 | 67.0 | 5.0 |
| H. Analgesic use at follow-up (binary outcome) | -1.25 | 0.21 | -0.25 (0.21) | 0.26 | 0.25 | 23.0 | 5.0 |

*the standard analytic method used in pain RCTs. LESI= lumbar epidural steroid injection, SE = standard error, Dep.= depression, Comp.= seeking disability

**Supplemental Table S10 (Scenario 3; also including covariates associated with analgesic use; $O_i \neq U_i$):** Mean estimates of beta coefficients (expected increase in observed pain intensity NRS outcome per unit increase in predictor variable), power to detect the treatment effect of LESI, and type I error, comparing different methods to account for analgesic use. Instead of having observed $O_i$ equal unobserved $U_i$ exactly for all patients $i = 1, \dots, n$ without analgesic use, $U_i$ was simulated to have 95-99% correlation with $O_i$ for these simulations.

| | Coefficient | | | | | | |
|---|---|---|---|---|---|---|---|
| **Analysis Method** | ***β₀*** **Intercept** | ***β₁*** **Baseline** | ***β₂*** **LESI (SE)** | ***β₃*** **Dep.** | ***β₄*** **Comp.** | **Power (%)** | **Type I error (%)** |
| Model values (as simulated) | 0.00 | 0.50 | -0.60 | 0.60 | 0.60 | | |
| Knowing underlying pain | 0.31 | 0.46 | -0.56 (0.20) | 0.57 | 0.56 | 78.7 | 5.2 |
| A. Ignoring analgesic use* | 0.15 | 0.37 | -0.45 (0.20) | 0.89 | 0.46 | 64.2 | 5.2 |
| B. Analgesic adjustment as a covariate | 0.08 | 0.36 | -0.43 (0.20) | 0.93 | 0.44 | 60.5 | 5.1 |
| C. Add 1.0 NRS points | 0.35 | 0.42 | -0.51 (0.21) | 0.75 | 0.51 | 70.9 | 5.4 |
| D. Add 1.5 NRS points | 0.45 | 0.45 | -0.54 (0.21) | 0.69 | 0.54 | 72.1 | 5.4 |
| E. Add 2.0 NRS points | 0.56 | 0.47 | -0.57 (0.23) | 0.62 | 0.57 | 72.4 | 5.4 |
| F. Exclude analgesic users | 0.13 | 0.36 | -0.43 (0.25) | 0.77 | 0.44 | 39.6 | 5.0 |
| G. Censored normal regression | 0.35 | 0.56 | -0.69 (0.29) | 0.38 | 0.71 | 68.3 | 5.5 |
| H. Analgesic use at follow-up (binary outcome) | -1.25 | 0.21 | -0.25 (0.21) | -0.58 | 0.25 | 23.1 | 5.1 |

*the standard analytic method used in pain RCTs. LESI= lumbar epidural steroid injection, SE = standard error, Dep.= depression, Comp.= seeking disability

**Supplemental Table S11 (Scenario 4; treatment affects analgesic use; $O_i \neq U_i$):** Mean estimates of beta coefficients (expected increase in observed pain intensity NRS outcome per unit increase in predictor variable), power to detect the treatment effect of LESI, and type I error, comparing different methods to account for analgesic use. Instead of having observed $O_i$ equal unobserved $U_i$ exactly for all patients $i = 1, \dots, n$ without analgesic use, $U_i$ was simulated to have 95-99% correlation with $O_i$ for these simulations.

| | Coefficient | | | | | | |
|---|---|---|---|---|---|---|---|
| **Analysis Method** | ***β₀*** **Intercept** | ***β₁*** **Baseline** | ***β₂*** **LESI (SE)** | ***β₃*** **Dep.** | ***β₄*** **Comp.** | **Power (%)** | **Type I error (%)** |
| Model values (as simulated) | 0.00 | 0.50 | -0.60 | 0.60 | 0.60 | | |
| Knowing underlying pain | 0.31 | 0.46 | -0.56 (0.20) | 0.57 | 0.56 | 78.7 | 5.2 |
| A. Ignoring analgesic use* | 0.12 | 0.38 | -0.33 (0.20) | 0.87 | 0.46 | 39.2 | 10.4 |
| B. Analgesic adjustment as a covariate | 0.05 | 0.36 | -0.28 (0.20) | 0.91 | 0.44 | 29.8 | 13.0 |
| C. Add 1.0 NRS points | 0.33 | 0.43 | -0.48 (0.21) | 0.74 | 0.52 | 65.8 | 5.6 |
| D. Add 1.5 NRS points | 0.44 | 0.45 | -0.56 (0.21) | 0.68 | 0.55 | 75.3 | 5.5 |
| E. Add 2.0 NRS points | 0.54 | 0.47 | -0.64 (0.23) | 0.61 | 0.57 | 81.5 | 6.4 |
| F. Exclude analgesic users | 0.08 | 0.36 | -0.28 (0.24) | 0.77 | 0.44 | 19.7 | 9.8 |
| G. Censored normal regression | 0.43 | 0.55 | -0.87 (0.27) | 0.42 | 0.68 | 89.6 | 12.6 |
| H. Analgesic use at follow-up (binary outcome) | -1.25 | 0.21 | -0.67 (0.21) | -0.58 | 0.24 | 89.2 | 51.9 |

*the standard analytic method used in pain RCTs. LESI= lumbar epidural steroid injection, SE = standard error, Dep.= depression, Comp.= seeking disability

**Supplemental Table S12: Sociodemographic and clinical characteristics among participants in the LESS RCT with available opioid analgesic data.**

| n (%) or Mean (Interquartile range) | Glucocorticoid -Lidocaine (N=169) | Lidocaine Alone (N=166) |
|---|---|---|
| Age | 67.7 (59, 75) | 67.7 (60, 75) |
| Female | 103 (61) | 85 (51) |
| Hispanic | 7 (4) | 6 (4) |
| Non-White Race | 55 (33) | 52 (31) |
| White Race | 114 (67) | 114 (69) |
| Married/living with partner | 104 (62) | 95 (57) |
| Employment status | | |
| *Full/part time* | 48 (28) | 58 (35) |
| *Retired, not disabled* | 82 (49) | 74 (45) |
| *Retired, disabled* | 24 (14) | 20 (12) |
| *Other* | 15 (9) | 14 (8) |
| Education | | |
| *High school equivalent or less* | 47 (28) | 57 (35) |
| *Some college, vocational/tech* | 63 (38) | 46 (28) |
| *College degree or more* | 58 (35) | 62 (38) |
| Duration of pain | | |
| *<3 months* | 21 (12) | 31 (19) |
| *3 to <12 months* | 52 (31) | 48 (29) |
| *1 to 5 years* | 53 (31) | 37 (22) |
| *>5 years* | 43 (25) | 49 (30) |
| Expectation of pain relief | 7.7 (7, 9) | 7.8 (6, 9) |
| Baseline Leg Pain intensity | 7.4 (6, 9) | 7.3 (6, 8) |
| Baseline Depression (PHQ-8)[a] | 7.3 (3, 11) | 6.1 (2, 8) |
| Baseline Anxiety (GAD-7)[b] | 4.7 (1, 7) | 4.8 (1, 6) |
| Baseline Fear Avoidance Beliefs Questionnaire | 19.9 (14, 26) | 19.0 (15, 26) |
| Baseline Pain Catastrophizing Scale | 18.6 (9, 26) | 18.1 (8, 26) |
| Baseline Quality of Life (EQ5D-VAS)[c] | 65.5 (50, 82) | 68.1 (60, 80) |

[a]PHQ-8: Patient Health Questionnaire depression 8-item scale
[b]GAD-7: Generalized Anxiety Disorder 7-item scale
[c]EQ5D-VAS: EuroQOL 5 dimensions visual analogue scale. Negative values for change in EQ5D-VAS indicates worse outcomes.
